# Submicroscopic malaria parasitemia and migration: the hidden weight carried by pregnant women

**DOI:** 10.1101/2024.01.21.24301565

**Authors:** L Docquier, C Taelman, M Delforge, C. Martin

## Abstract

**Introduction:** Submicroscopic malaria parasitemia is a known phenomenon for years, however there are currently not enough existing data to set guidelines for diagnosis and treatment. Due to multiple reasons, pregnant women arrived recently from endemic areas seem to be particularly at risk of submicroscopic malaria parasitemia, which can only be diagnosed by molecular techniques. This study aims to determine the prevalence of submicroscopic malaria parasitemia in pregnant women coming from endemic areas and to pin down its potential consequences on maternal and birth outcomes.

**Methods:** A prospective study on pregnant women that recently arrived from endemic areas for malaria was conducted in the Obstetrics Department of CHU Saint-Pierre (Brussels). Several *P. falciparum* detection techniques were performed for each participant on 2 maternal (pre and postnatal) and 1 cord blood samples, using both conventional tests and loop-mediated isothermal amplification (LAMP). Placentas were also analyzed to search for placental malaria. Birth outcomes were collected. Patients with and without submicroscopic malaria parasitemia were compared.

**Results:** Maternal and umbilical blood samples were collected in 96 enrolled patients and analyzed by conventional tests and loop mediated isothermal amplification. Among the enrolled patients, 5 (5.2%) had submicroscopic malaria parasitemia. Patients with submicroscopic malaria parasitemia had left their endemic area more recently (p <0.05), had higher reticulocyte counts in both maternal prenatal blood (p <0.02) and cord blood (p <0.02) and had higher lactate dehydrogenase level in cord blood (p <0.05). No impact of submicroscopic malaria parasitemia on birth weight and term of birth was detected.

**Conclusion:** The prevalence of submicroscopic malaria parasitemia in pregnant women arriving in non-endemic countries is non negligible. More data need to be collected to assess the consequences of submicroscopic malaria parasitemia on maternal and birth outcomes.

## Introduction

### 1.1. Context and scientific motivation

Nowadays, *Plasmodium falciparum* (*P. falciparum*) malaria in a non-immunized subject (known as primary invasion) is well known (1). There are international guidelines for its diagnosis, treatment and prevention, since it rapidly presents an immediate vital risk for the person who develops it (2).

However, repeated episodes of malaria in patients living in highly endemic zones (API (Annual Parasite Index) >100), through chronic stimulation of the immune system, allow patients to develop a partial, mostly humoral immunity which protects them from symptoms while carrying the parasite: it is called “premunition” (3). In endemic areas, this phenomenon may lead to a persistent parasitemia that oscillates with exposure and time and causes few or no clinical symptoms (4).

This low-density infection in a person benefiting from premunition is hardly detectable with conventional diagnostic methods (thin smear, thick drop, rapid diagnostic test (RDT)) and can only be detected with the development of more recent screening techniques (3). Loop-mediated isothermal amplification (LAMP), a technique almost as sensitive as the polymerase chain reaction (PCR), but less expensive, more accessible, and easier in a technical point of view, is able to detect submicroscopic malaria (5).

As this submicroscopic malaria parasitemia is not systematically diagnosed, the prevalence and consequences of this phenomenon still need to be determined (6). However, it is known that if humoral immunity decreases for one reason or another, parasitemia can increase and lead to actual symptomatic, possibly severe malaria. These reasons are, for example: immunosuppression, pregnancy, or departure from the endemic zone with the absence of the repeated stimuli that maintain humoral immunity (7).

Migrants coming from malaria endemic areas are likely to carry submicroscopic malaria parasitemia for a variable period of time (8) and to slowly lose the protection conferred by repeated contact with the parasite. It is thought that the premunition disappears 12-24 months after leaving the endemic area (9). It therefore exposes migrants to malaria reactivation and sometimes to the risk of severe malaria (10).

### 1.2. State of the art and objectives

For several reasons, pregnant women are particularly vulnerable to *P. falciparum* infections and its complications (11):

- Due to hormonal and immunological changes, pregnant women are more susceptible to malaria infection, especially during the first pregnancy (4)
- Because of the decrease in cellular immunity during pregnancy, pregnant women are at increased risk to present severe malaria episodes (12)
- *P. falciparum* has a particular affinity for the placenta, where it gets sequestered by variant surface antigen 2 chondroitin sulfate A (VAR2CSA) receptors. As a result, the blood parasitemia is low in pregnant women, hampering the malaria diagnosis with the conventional methods (13).

Consequently, pregnant women who have recently arrived from endemic areas appear to be a group at risk for submicroscopic malaria parasitemia, which can lead to severe reactivation (10). Depending on the prevalence and the consequences of that infection (both still to be determined), these women (and offspring) might benefit from early detection and treatment upon arrival in non-endemic countries.

For the first time in a non-endemic country, we aimed to determine the prevalence of submicroscopic malaria parasitemia (defined as thin smear -, thick drop -, RDT -, LAMP +) in a sample of pregnant women arriving from endemic areas. We also aimed to assess the consequences of this submicroscopic malaria parasitemia on the biological parameters of the mother and the offspring, the term of delivery and the weight at birth.

## Methods

### Recruitment and population

From January 2020 until July 2022, a prospective study was conducted at the Centre Hospitalier Universitaire (CHU) Saint-Pierre, a large tertiary public hospital in the center of Brussels where a large majority of patients come from different countries. In the prenatal consultations, 16.5% of the followed patients are from sub-Saharan Africa.

During prenatal consultations, pregnant migrant women arriving from endemic areas for malaria (2) were recruited. Inclusion criteria were age >18 years old and date of departure of the endemic zone (not more than 2 years before the beginning of their pregnancy). Patients with malaria symptoms or having received antimalarial treatment during the current pregnancy were excluded from recruitment.

The patient’s electronic health record (EHR) was used to set eligibility. During any of the antenatal appointments, the study was explained to the patient. If she accepted to participate, the study informed consent form (ICF) was signed, and a first blood sample was taken. During delivery, a second blood sample was taken from the umbilical cord and the placenta was sent to the Pathological Anatomy Laboratory. On day 1 postpartum, a third blood sample was taken from the mother in the maternity ward.

(see Figure 1)

**Fig. 1:**
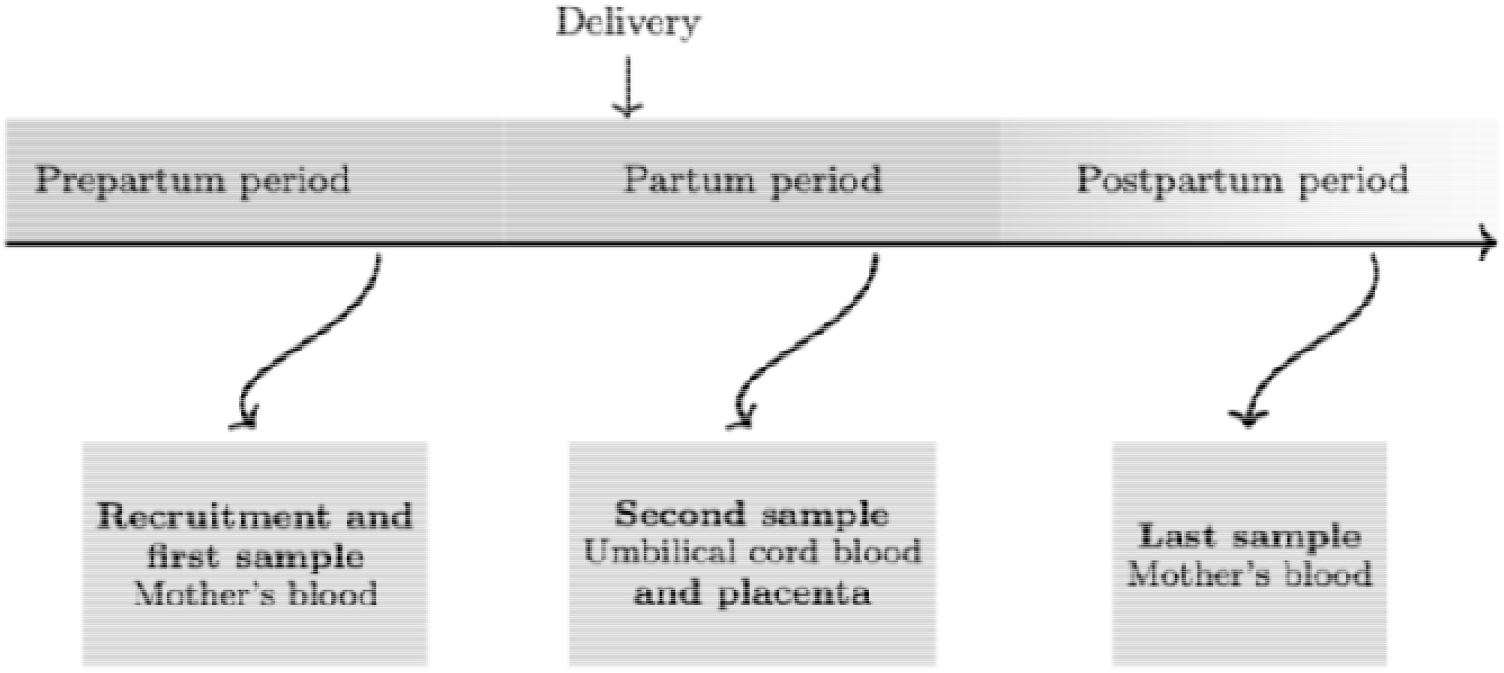
Patient’s recruitment and follow-up.

Other information was obtained from the EHR, including HIV status and presence of sickle cell disease. At any time, if microscopic malaria was diagnosed, an antimalarial treatment was proposed. If submicroscopic malaria was diagnosed, no treatment was proposed.

### Laboratory techniques

Blood samples were all analyzed at the Laboratoire Hospitalier Universitaire de Bruxelles - Universitair Laboratorium van Brussel (LHUB-ULB). Biomarkers of hemolytic anemia were searched for (hemoglobin, reticulocytes, lactate dehydrogenase (LDH), haptoglobin, bilirubin) and the presence of *P. falciparum* was determined both with routine molecular tests (thin smear, thick drop) and antigen-based tests (the Illumigene Malaria® test using LAMP technique). The placentas were sent to the Pathological Anatomy Laboratory of the Bordet Institute (Brussels) where they were fixed and analyzed by a specialized anatomopathologist. Macroscopic and microscopic analyses of the membranes, cord and placenta were carried out to search for a placental malaria, defined by an infiltration of the intervillous spaces by parasitized red blood cells, inflammatory cells and hemozoin deposits. Any other placental anomalies noted were also recorded.

### Pregnancy outcomes

Information regarding the term of delivery, occurrence of (pre-)eclampsia, birth weight, and fetal death were collected using the EHR.

### Data Analysis and statistical methods

Collected data were anonymized then manually entered in a REDCapTM database. REDCap (Research Electronic Data Capture) is a secure, web-based software platform designed to support data capture for research studies (14). Descriptive statistics were used to summarize the characteristics of our population and groups. Two different analyses were made to assess submicroscopic malaria potential risk factors or consequences. The first one strictly compared LAMP+ to LAMP-samples for each kind of sample. The second one compared patients with at least one positive LAMP test during the follow-up to patients without. Hypothesis tests for differences between groups were performed using non-parametric Wilcoxon-Mann-Whitney tests for continuous variables, and Fisher exact tests for categorical variables. We used SAS statistical software (version 9.4 SAS institute, Cary, North Carolina, USA) for all statistical analyses. All p-values were 2-sided and considered statistically significant if p <0.05.

### Ethics Committee

This study was approved by the Ethics Committee of the Centre Hospitalier Universitaire (CHU) Saint-Pierre (CE/19-10-12).

## Results

A total of 96 patients were enrolled from January 2020 to July 2022: 95 prepartum blood samples were collected, 72 cord blood samples, 75 (day 1) postpartum blood samples (totaling 242 blood samples) and 56 placentas were available for analysis. No microscopic malaria was diagnosed by thin or thick drop.

### Prevalence of submicroscopic malaria

Due to hemolyzed samples, 236 on the 242 samples were analyzed for *P. falciparum*. Among them, 7 samples (7/236, 2.97%) were positive for submicroscopic malaria (Table 1) in 5 of the 96 included patients (5/96, 5.2%).

**Table 1.** Malaria detection tests.

| Test | Positive |
| --- | --- |
| **RDT** |  |
| Prepartum | 0/94 (0%) |
| Cord blood | 0/67 (0%) |
| Postpartum | 0/75 (0%) |
| **Thick drop** |  |
| Prepartum | 0/94 (0%) |
| Cord blood | 0/67 (0%) |
| Postpartum | 0/75 (0%) |
| **LAMP** |  |
| Prepartum | 4/93 (4.3%) |
| Cord blood | 1/67 (1.49%) |
| Postpartum | 2/75 (2.67%) |

*LAMP: loop-mediated isothermal amplification; RDT: rapid diagnostic test*

A summary of the patients with at least one positive LAMP test is presented in Table 2.

**Table 2.**
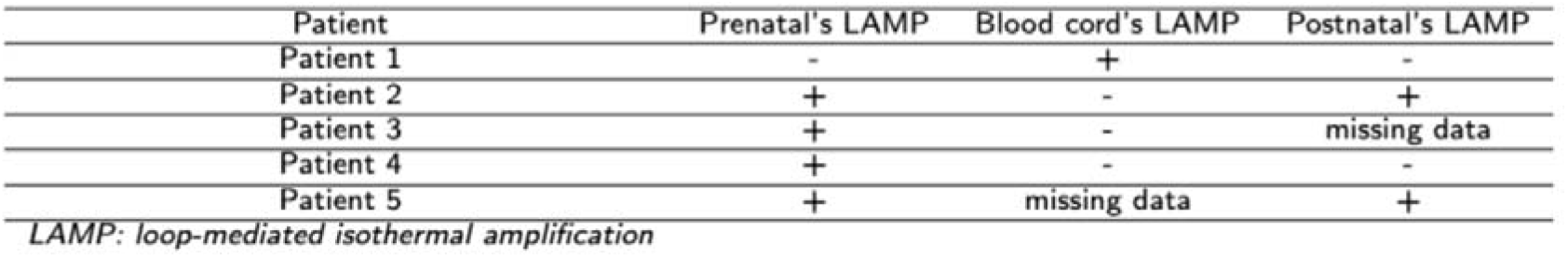
Summary of patients with at least one positive LAMP test.

### Socio demographic characteristics of the study population

The 96 patients were born in 16 different countries following the distribution shown in Figure 2. A majority were coming from West Africa and Central Africa with the 3 most represented countries of origin being Guinea (29.17%), Cameroon (19.79%) and Democratic Republic of Congo (13.54%). The countries of birth of patients that had at least one positive LAMP test are highlighted with circles on the same figure (three on the five patients were coming from Cameroon, one from Ivory Coast and the last from Togo).

**Fig. 2:**
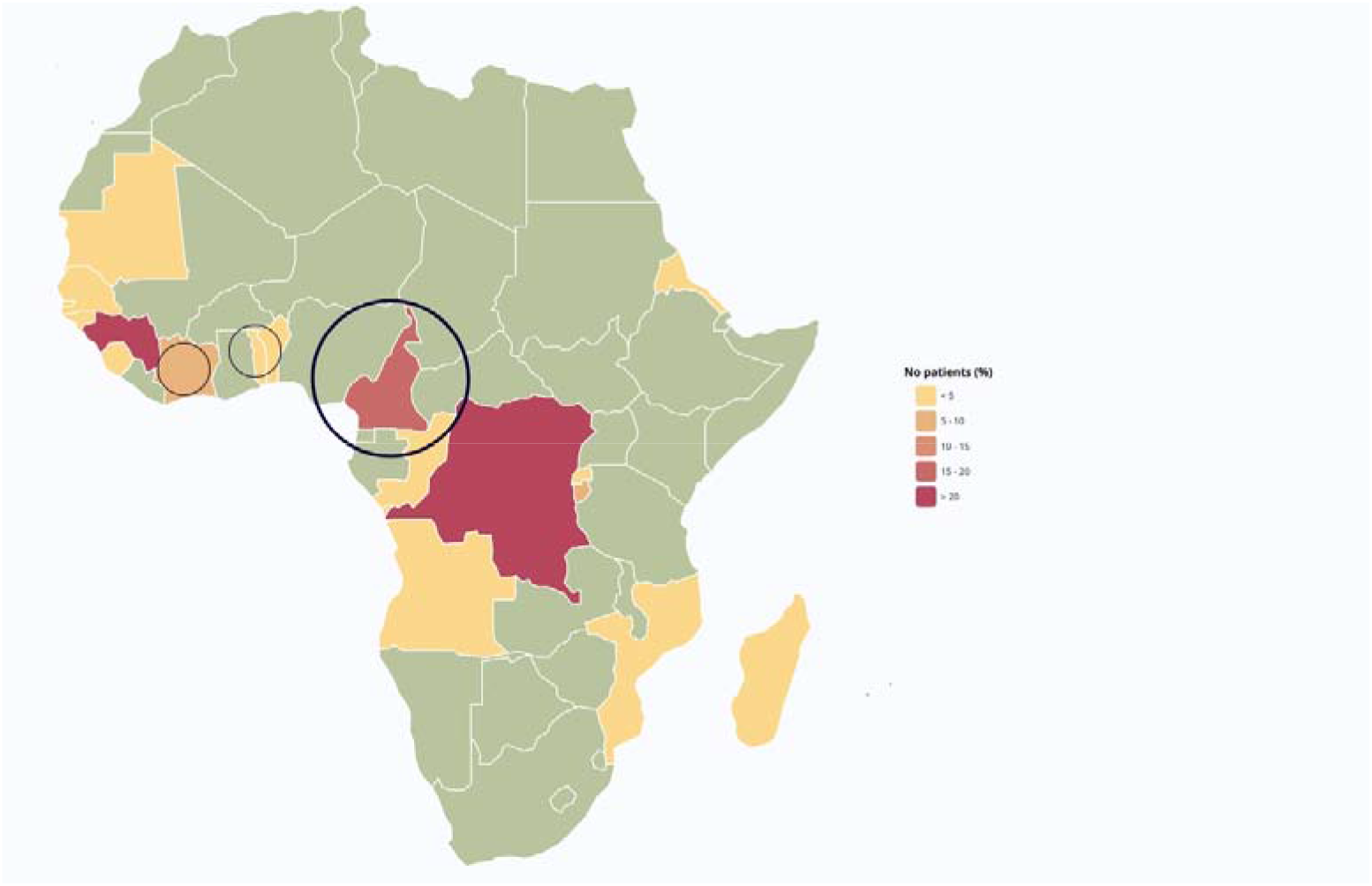
Countries of birth of enrolled patients.

Socio-demographic characteristics of the enrolled patient and comparison between patients with at least one positive LAMP test during the follow-up and the others are presented in Table 3.

**Table 3.** Study population characteristics at inclusion.

| Characteristics | All (n= 96) | LAMP- (n= 91) | LAMP+ (n= 5) | p-value |
| --- | --- | --- | --- | --- |
| **Age, median (IQR), years** | 29.5 (25.7-32.3) | 29.2 (25.5-32.2) | 31.3 (31.1-35.2) | 0.06 |
| **Parity, no (%)** |  |  |  | 1 |
| Primiparous | 30 (31.25) | 29 (31.87) | 1 (20) |  |
| Multiparous | 66 (68.75) | 62 (68.13) | 4 (80) |  |
| **Gestational age, median (IQR), weeks** | 27 (22-32.5) | 27 (22-33) | 29 (22-30) | 0.96 |
| **Foetuses, no (%)** |  |  |  | 0.68 |
| Single pregnancy | 89 (92.71) | 84 (92.31) | 5 (100) |  |
| Twin pregnancy | 6 (6.25) | 6 (6.59) | 0 (0) |  |
| Triplet pregnancy | 1 (1.04) | 1 (1.1) | 0 (0) |  |
| **Environment, no (%)** |  |  |  | 0.16 |
| Urban area | 56 (58.33) | 52 (57.14) | 4 (80) |  |
| Rural region | 7 (7.29) | 6 (6.59) | 1 (20) |  |
| Unknown | 33 (34.38) | 33 (36.26) | 0 (0) |  |
| **Endemicity, no (%)** |  |  |  | 1 |
| Moderately endemic area | 9 (9.37) | 9 (9.89) | 0 (0) |  |
| Highly endemic area | 87 (90.62) | 82 (90.11) | 5 (100) |  |
| **Duration of stay in Europe, median (IQR), months** | 9.4 (5.4-17.9) | 9.6 (5.4-18) | 7.9 (7.9-10.5) | 0.55 |
| **Duration of stay in Europe, no (%)** |  |  |  | 0.04 |
| <6 months | 29 (30.21) | 28 (30.77) | 1 (20) |  |
| 6-12 months | 30 (31.25) | 26 (28.57) | 4 (80) |  |
| > 12 months | 37 (38.54) | 37 (40.66) | 0 (0) |  |
| **Comorbidities, no (%)** |  |  |  |  |
| **HIV+** | 6 (6.32) | 5 (5.49) | 1 (20) | 0.28 |
| **Sickle cell disease** |  |  |  | 1 |
| AC | 1 (1.08) | 1 (1.12) | 0 (0) |  |
| AS | 16 (17.2) | 15 (16.85) | 1 (20) |  |
| SS | 1 (1.08) | 1 (1.12) | 0 (0) |  |
| **HBV+** | 7 (7.51) | 6 (6.82) | 1 (20) | 0.37 |
| **Diabetes** | 18 (18.75) | 18 (19.78) | 0 (0) | 0.58 |
| **History of Malaria, no (%)** | 75 (78.95) | 71 (78.89) | 4 (80) | 1 |
| **Number of episodes, median (IQR)** | 6.5 (4-13.5) | 6.5 (4-13.5) | 7.5 (5-17.5) | 0.60 |

*HBV: hepatitis B virus; HIV: human immunodeficiency virus; IQR: interquartile range; LAMP: loop-mediated isothermal amplification; no: number*

The median gestational age at sampling was 27 weeks (IQR: 22-32.5). All patients with submicroscopic malaria parasitemia (5/5) were coming from 3 highly endemic areas (circled down on Fig 2) and left it less than one year before inclusion. Four of the five patients with submicroscopic malaria parasitemia were multiparous and one of the five was HIV-positive.

### Hematological parameters in relation to submicroscopic *P. falciparum* infection

Hematological parameters in the prenatal, cord and postpartum blood samples as well as birth outcomes are shown in Table 4.

**Table 4.** Hematological parameters in the prenatal sample, the cord blood sample and the postpartum sample.

| Hematological parameters | All (n= 96) | LAMP- (n= 91) | LAMP+ (n= 5) | p-value |
| --- | --- | --- | --- | --- |
| **Hemoglobin**, median (IQR), g/dl |  |  |  |  |
| Prepartum sample | 11.2 (10.5-11.7) | 11.2 (10.5-11.7) | 11.3 (11.1-11.8) | 0.56 |
| Cord blood sample | 14.9 (13.9-16) | 14.9 (14-16.2) | 14.7 (14.4-16.1) | 0.78 |
| Postpartum sample | 10.4 (9.5-11.6) | 10.4 (9.5-11.6) | 10.1 (8.7-10.6) | 0.35 |
| **Reticulocytes** |  |  |  |  |
| Median (IQR), % |  |  |  |  |
| Prepartum sample | 2.2 (1.8-2.6) | 2.2 (1.8-2.5) | 2.7 (2.7-3) | <0.05 |
| Cord blood sample | 4.1 (3.6-4.7) | 4 (3.5-4.6) | 5.2 (5-5.9) | <0.02 |
| Postpartum sample | 2.3 (2.1-2.8) | 2.3 (2.1-2.8) | 2.6 (2.2-3.2) | 0.48 |
| Median (IQR), 10<sup>3</sup>/μL |  |  |  |  |
| Prepartum sample | 83.5 (72.3-96.8) | 81.8 (72.2-95.3) | 102.1 (96.8-109.8) | 0.02 |
| Cord blood sample | 173 (151.5-199) | 170.6 (150.3-195.3) | 222.6 (209.4-248) | <0.01 |
| Postpartum sample | 82.8 (74-98.7) | 82 (74-98) | 93.9 (72.3-106.9) | 0.68 |
| **LDH**, median (IQR), IU/L |  |  |  |  |
| Prepartum sample | 216.5 (199-242) | 217 (198-239) | 215 (214-249) | 0.70 |
| Cord blood sample | 488 (423-549) | 482 (423-546) | 596 (558-642.5) | 0.04 |
| Postpartum sample | 309 (283-353) | 310 (283-354) | 285 (259-288) | 0.07 |
| **Haptoglobin**, median (IQR), mg/dL |  |  |  |  |
| Prepartum sample | 86 (55-120) | 86 (54-121) | 100 (58-118) | 0.82 |
| Cord blood sample | 10 (10-10) | 10 (10-10) | 10 (10-10) | 1 |
| Postpartum sample | 101 (70-130) | 101 (68-129) | 121 (92.5-153.5) | 0.30 |
| **Bilirubin**, median (IQR), mg/dL |  |  |  |  |
| Conjugated |  |  |  |  |
| Prepartum sample | 0.1 (0.1-0.1) | 0.1 (0.1-0.1) | 0.1 (0.1-0.1) | 0.99 |
| Cord blood sample | 0.3 (0.2-0.4) | 0.3 (0.2-0.4) | 0.3 (0.3-0.3) | 1 |
| Postpartum sample | 0.1 (0.1-0.2) | 0.1 (0.1-0.2) | 0.2 (0.1-0.2) | 0.55 |
| Total |  |  |  |  |
| Prepartum sample | 0.2 (0.2-0.3) | 0.2 (0.2-0.3) | 0.2 (0.2-0.4) | 0.81 |
| Cord blood sample | 1.6 (1.3-1.9) | 1.6 (1.4-1.9) | 1.9 (1.7-2.4) | 0.10 |
| Postpartum sample | 0.3 (0.2-0.4) | 0.3 (0.2-0.5) | 0.3 (0.2-0.4) | 0.81 |
| **Platelets**, median (IQR), 10<sup>3</sup>/μL |  |  |  |  |
| Prepartum sample | 228 (187-256) | 228 (189-257) | 184 (181-229) | 0.19 |
| Cord blood sample | 280 (225-314) | 284 (225-316) | 240 (224.5-251.5) | 0.13 |
| Postpartum sample | 188 (161-226) | 188 (160-226) | 188.5 (173.5-192.5) | 0.76 |

IQR: interquantile range; LAMP: loop-mediated isothermal amplification; LDH: lactate dehydrogenase

#### Prenatal Period

4/95 (4.21%) prenatal blood samples were positive in LAMP. Comparison between patients that had at least one positive LAMP test during the follow-up and the others showed a higher reticulocyte level in patients with submicroscopic malaria (p <0.05 for relative and p= 0.02 for absolute reticulocyte counts).

#### Delivery period

##### Cord blood samples

72 cord blood samples were sent to the laboratory, one was positive for LAMP (1/72, 1.39%). Cord blood of the patients with at least one positive LAMP during the follow-up had a higher reticulocyte level (p <0.02 for relative and p <0.01 for absolute reticulocyte counts) and a higher LDH level (p= 0.04) than what was measured in the cord blood of patients without submicroscopic malaria parasitemia.

#### Placentas

56 placentas were analyzed, and none showed signs of placental malaria. 51 placentas (91%) showed other abnormalities (see Table 5) whereas none was evocative of malaria in pregnancy.

**Table 5:** placentas abnormalities (n=51)

| Placentas abnormalities (n= 51) |  |  |  |
| --- | --- | --- | --- |
| Type of abnormality | No | Type of abnormality | No |
| Trophoblastic cyst | 12 | Intravascular thrombosis | 3 |
| Chronic deciduitis | 11 | Decidual arteriopathy | 3 |
| Old infarct | 10 | Sickle cell stigmata | 3 |
| Intervillous haemorrhage | 11 | Chronic Lymphocytic chorioamnionitis | 2 |
| Villous hypoxia-ischemia | 14 | Intervillous thrombosis | 5 |
| Villous ischemic necrosis | 8 | Chronic villitis | 1 |
| Congestive maternal circulation | 4 | Subacute deciduitis | 1 |
| Acute amnionitis | 6 | Chorionic cyst | 1 |
| Acute chorioamnionitis | 4 | Chronic intervillitis | 1 |
| Congestive foetal circulation | 1 | Villous immaturity | 1 |

There was no significant difference between the placentas of patients with and without submicroscopic malaria parasitemia (p> 0.5).

#### Birth outcomes

96 babies were born from 88 mothers. Four mothers (4.5%), all of them LAMP-, suffered from pre-eclampsia.

Between patients with at least one positive LAMP during the follow up and the others, there was no significant difference in birth outcomes (see Table 6).

**Table 6:**
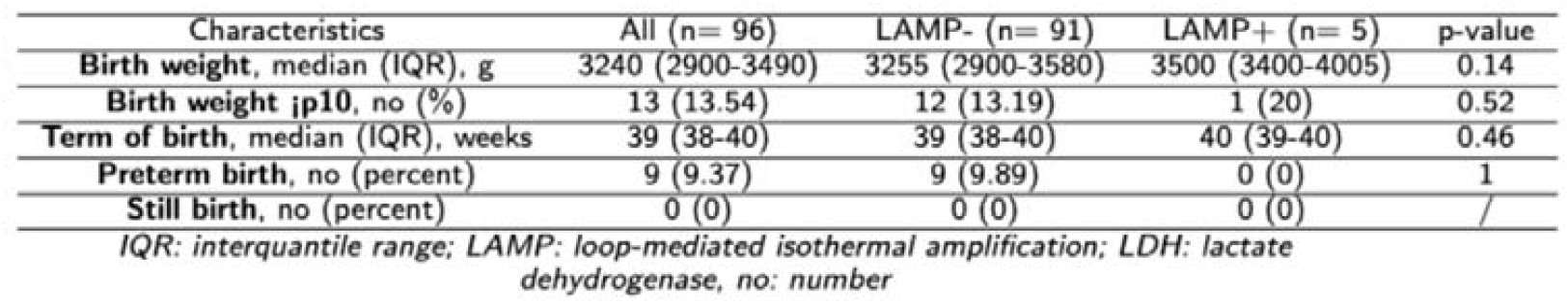
Birth outcomes.

#### Postpartum period

2/75 (2.67 %) maternal postpartum blood tests were positive for LAMP. There was no significant difference in hematological parameters between patients with at least one positive LAMP test during the follow-up and the others.

## Discussion

The prevalence of submicroscopic malaria parasitemia was 5.2% (5/95) in the included patients. All patients with at least one positive LAMP test (n= 5) were coming from 3 malaria hyper-endemic (API> 100) countries (Cameroon, Ivory Coast, Togo). Higher prevalence of submicroscopic malaria parasitemia in hyper-endemic countries (compared to what we can find in low malaria transmission areas) can be explained by acquired immunity due to exposure: individuals who are frequently exposed to Plasmodium infections over time are more likely to develop partial anti-disease immunity and thus, to carry submicroscopic malaria (15).

While Malaria in Pregnancy in endemic areas is held responsible for 11% of all newborn deaths (16) and increases risk of preterm delivery, low birth weight, maternal and fetal anemia (17), the prevalence and consequences of submicroscopic malaria in pregnant women remain to be determined. In hyper-endemic areas, prevalence of submicroscopic malaria parasitemia is shown to be twice more frequent in the general population than microscopic malaria (63.7% versus 31.2%) (18). In pregnant women, in all endemic areas, a systematic review estimated a prevalence of submicroscopic malaria parasitemia of 13.5% (more common than microscopic malaria parasitemia which has a prevalence of 8% in this meta-analysis) (19).

Studies have tried to assess the consequences of this submicroscopic malaria, but results were discrepant: while one study in Benin showed significantly increased risk of maternal anemia, of low birth weight in primigravidae and of premature births in multigravidae (20), two other studies in Malawi (6) (21) showed no association between submicroscopic malaria infection and adverse delivery outcomes. However, differences in definitions, power of analyses and country endemicity make the results difficult to compare.

In this context, performing a first prospective study about submicroscopic malaria in pregnant women outside endemic areas is important for multiple reasons. First, results of studies in endemic regions are discrepant since each country has its own endemicity, its own national treatment guidelines (intermittent preventive treatment being different following the country) and its own confounding biases. Then, migrating pregnant women are more at risk of reactivation and migration from sub-Saharan Africa has increased in Belgium these last few years (22). Finally, pregnant women stop taking intermittent preventive treatment for malaria when arriving in Europe, when they could carry the parasite up to 24 months (9).

In this study, the 5 patients with submicroscopic malaria parasitemia had left the highly endemic area more recently (between 6 and 12 months before inclusion, p <0.05) than the patients without submicroscopic malaria parasitemia.

There was no difference in parity observed between patients with and without submicroscopic malaria parasitemia. In the literature, increased parity is thought to protect against symptoms of malaria in endemic areas. Indeed, as *P. falciparum* is sequestrated by binding receptors (VAR2CSA) in the placenta, multiparous women can develop anti-adhesion antibodies protecting them from *Plasmodium* infection in subsequent pregnancies (15), decreasing the risk of microscopic malaria in favor of submicroscopic malaria parasitemia which is shown to be more frequent in multigravid women and older women (>30 years) than in primigravid women and younger women (19).

In this cohort, one (of the 5 patients with submicroscopic malaria parasitemia) woman was HIV-positive but we found no statically significant difference between patients with and without submicroscopic malaria parasitemia regarding HIV status even if HIV coinfection increases prevalence, frequency, duration and/or intensity of Plasmodium carriage (23).

If these elements are confirmed in large-scaled studies and if deleterious consequences are identified, pregnant women to screen as a priority for submicroscopic malaria could be targeted on the basis of the country of origin endemicity (API >100), the date of departure from the endemic area (less than one year before the first prenatal care appointment), the parity (nulliparous women being more at risk of malaria complications) and the HIV status (HIV-positive patients being more vulnerable to *P. falciparum* carriage).

In this study, there was no difference in prevalence of anemia between patients with and without submicroscopic malaria parasitemia, despite the association evoked in the literature (24). However, reticulocyte levels were significantly higher in patients with submicroscopic malaria both in the prenatal maternal blood and in the cord blood. As hemolysis linked to malaria classically causes a rise of reticulocytosis, there is a theorical plausibility that a subpatent malaria infection could cause a chronic and low-grade hemolysis without interfering with hemoglobin levels yet. In the same way, LDH levels were significantly higher in the cord blood of the babies born from patients with submicroscopic malaria, accordingly to the literature: as *P. falciparum* classically uses LDH for its glycolytic cycle, LDH is overexpressed in malaria infection (24).

These two observations highlight that submicroscopic malaria has consequences on the red blood cell cycle. It can therefore be hypothesized that if the study was to be scaled up, consequences on maternal/foetal outcomes could be observed.

At this stage, we were not able to highlight any birth adverse outcome (in terms of low birth weight and preterm) linked to submicroscopic malaria: this could be explained by a lack of statistical power due to the size of the sample (n= 96) and/or the prevalence of submicroscopic malaria in our sample (5.2%) or by an absence of association between submicroscopic malaria and adverse birth outcomes. The main strength of this study is its longitudinal prospective design exploring a subject never investigated before outside endemic areas.

Its limitations are the missing data, the relatively low number of included patients and the short follow-up time. Are there any sequalae of this subpatent infection over time on the offspring’s immunological system, on its growth curve, etc.? As chronic infections are defined by an altered transcriptional profile and by a persistent inflammation that influences the immune system (25), immunology effects of submicroscopic malaria in mothers and babies still need to be determined.

## Conclusion

Submicroscopic malaria is present in a small although non negligible proportion of pregnant women arriving from endemic areas. These first results in a non-endemic area need to be confirmed in large-scaled studies to assess potential consequences of this infection.

## Supporting information

cover letter

## Data Availability

All data produced in the present study are available upon reasonable request to the authors

## References

1. Plewes K, Leopold SJ, Kingston HGF, Dondorp AM. Malaria: What’s New in the Management of Malaria? Infect Dis Clin North Am. 2019; 33(1):39–60. doi: 10.1016/j.idc.2018.10.002.

2. Word Health Organization [Online]. Geneva: WHO;2022. WHO guidelines for malaria. Available from: https://www.who.int/publications/i/item/guidelines-for-malaria [accessed 2nd April 2022].

3. Nguyen TN, von Seidlein L, Nguyen TV, Truong PN, Do Hung S, Pham HT et al. The persistence and oscillations of submicroscopic Plasmodium falciparum and Plasmodium vivax infections over time in Vietnam: an open cohort study. Lancet Infect Dis. 2018; 18 (5):565–72. doi: 10.1016/S1473-3099(18)30046-X.

4. Laishram DD, Sutton PL, Nanda N, Sharma VL, Sobti RC, Carlton JM et al. The complexities of malaria disease manifestations with a focus on asymptomatic malaria. Malar J. 2012;11:29. doi:10.1186/1475-2875-11-29.

5. Feleke DG, Alemu Y, Yemanebirhane N. Performance of rapid diagnostic tests, microscopy, loop-mediated isothermal amplification (LAMP) and PCR for malaria diagnosis in Ethiopia: a systematic review and meta-analysis. Malar J. 2021; 20:384. doi: 10.1186/s12936-021-03923-8.

6. Cohee LM, Kalilani-Phiri L, Boudova S, Joshi S, Mukadam R, Seydel KB et al. Submicroscopic malaria infection during pregnancy and the impact of intermittent preventive treatment. Malar J. 2014;13:274. doi: 10.1186/1475-2875-13-274.

7. Ndam NT, Mbuba E, González R, Cisteró P, Kariuki S, Sevene E et al. Resisting and tolerating P. falciparum in pregnancy under different malaria transmission intensities. BMC Med. 2017;15:130. doi: 10.1186/s12916-017-0893-6.

8. Monge-Maillo B, Norman F, Pérez-Molina JA, Díaz-Menéndez M, Rubio JM, López-Vélez R. Plasmodium falciparum in Asymptomatic Immigrants from Sub-Saharan African, Spain. Emerg Infect Dis. 2012;18(2):356–7. doi:10.3201/eid1802.111283.

9. Lindblade KA, Steinhardt L, Samuels A, Kachur SP, Slutsker L. The silent threat: asymptomatic parasitemia and malaria transmission. Expert Rev Anti Infect Ther. 2013;11(6):623–39. doi: 10.1586/eri.13.45.

10. Askling HH, Bruneel F, Burchard G, Castelli F, Chiodini PL, Grobusch MP et al. Management of imported malaria in Europe. Malar J. 2012;11:328. doi: 10.1186/1475-2875-11-328.

11. Desai M, ter Kuile FO, Nosten F, McGready R, Asamoa K, Brabin B et al. Epidemiology and burden of malaria in pregnancy. Lancet Infect Dis. 2007;7(2):93–104. doi: 10.1016/S1473-3099(07)70021-X.

12. Manirakiza A, Serdouma E, Ngbalé RN, Moussa S, Gondjé S, Degana RM et al. A brief review on features of falciparum malaria during pregnancy. J Public Health Afr. 2017;8(2):668. doi: 10.4081/jphia.2017.668.

13. Rogerson SJ, Hviid L, Duffy PE, Leke RF, Taylor DW. Malaria in pregnancy: pathogenesis and immunity. Lancet Infect Dis. 2007;7(2):105–17. doi: 10.1016/S1473-3099(07)70022-1.

14. Harris PA, Taylor R, Minor BL, Elliott V, Fernandez M, O’Neal L et al. The REDCap consortium: Building an international community of software platform partners. J Biomed Inform. 2019;95:103208. doi: 10.1016/j.jbi.2019.103208.

15. Yimam Y, Nateghpour M, Mohebali M, Abbaszadeh Afshar MJ. A systematic review and meta-analysis of asymptomatic malaria infection in pregnant women in Sub-Saharan Africa: A challenge for malaria elimination efforts. PLoS One. 2021;16(4):e0248245. doi: 10.1371/journal.pone.0248245.

16. Bakken L, Iversen PO. The impact of malaria during pregnancy on low birth weight in East-Africa: a topical review. Malar J. 2021;20:348. doi: 10.1186/s12936-021-03883-z.

17. Mahamar A, Andemel N, Swihart B, Sidibe Y, Gaoussou S, Barry A et al. Malaria Infection Is Common and Associated With Perinatal Mortality and Preterm Delivery Despite Widespread Use of Chemoprevention in Mali: An Observational Study 2010 to 2014. Clin Infect Dis. 2021;73(8):1355–61. doi: 10.1093/cid/ciab301.

18. Mbama Ntabi JD, Lissom A, Djontu JC, et al. Prevalence of non-Plasmodium falciparum species in southern districts of Brazzaville in The Republic of the Congo. Parasit Vectors. 2022;15(1):209. doi:10.1186/s13071-022-05312-9

19. van Eijk AM, Stepniewska K, Hill J, et al. Prevalence of and risk factors for microscopic and submicroscopic malaria infections in pregnancy: a systematic review and meta-analysis. Lancet Glob Health. 2023;11(7):e1061–e1074. doi:10.1016/S2214-109X(23)00194-8.

20. Cottrell G, Moussiliou A, Luty A, Cot M, Fievet N, Massougbodji A et al. Submicroscopic Plasmodium falciparum Infections Are Associated With Maternal Anemia, Premature Births, and Low Birth Weight. Clin Infect Dis. 2015; 60(10):1481–8. doi: 10.1093/cid/civ122.

21. Taylor SM, Madanitsa M, Thwai KL, Khairallah C, Kalilani-Phiri L, van Eijk AM et al. Minimal Impact by Antenatal Subpatent Plasmodium falciparum Infections on Delivery Outcomes in Malawian Women: A Cohort Study. J Infect Dis. 2017;216(3):296–304. doi: 10.1093/infdis/jix304.

22. Centre Fédéral Migration [Online]. Myria;2021. La Migration en Chiffres et en Droits : Cahier “Population et Mouvements”. Available from: https://www.myria.be/fr/publications/le-rapport-migration-2021-sous-forme-de-cahiers [accessed 2nd June 2023].

23. Roberds A, Ferraro E, Luckhart S, Stewart VA. HIV-1 Impact on Malaria Transmission: A Complex and Relevant Global Health Concern. Front Cell Infect Microbiol. 2021;11:656938. doi: 10.3389/fcimb.2021.656938.

24. Jain P, Chakma B, Patra S, Goswami P. Potential biomarkers and their applications for rapid and reliable detection of malaria. Biomed Res Int. 2014;2014:852645. doi: 10.1155/2014/852645.

25. Álvarez Larrotta C, Agudelo OM, Duque Y, Gavina K, Yanow SK, Maestre A et al. Submicroscopic Plasmodium infection during pregnancy is associated with reduced antibody levels to tetanus toxoid. Clin Exp Immunol. 2019;195(1):96–108. doi: 10.1111/cei.13213.

