## Supplementary material for "Submicroscopic malaria parasitemia and migration: the hidden weight carried by pregnant women": cover letter

Léa Docquier^1^, Chloë Taelman^1^, Martin Delforge and Charlotte Martin

^1^These first authors contributed equally to this article.

All co-authors have agreed to the submission.

*Author’s contributions:*
Docquier L and Taelman C enrolled patients and wrote the manuscript. Both DL and TC contributed to the final version of this project and are designated as co-first authors.
Delforge M performed the statistics.
Martin C conceived the original idea and supervised the project.

*Author affiliations:*

Université Libre de Bruxelles, Brussels, Belgium (L. Docquier, C. Taelman)

Centre Hospitalier Universitaire Saint-Pierre, Brussels, Belgium (M. Delforge, C. Martin)

**Article Summary Line**

The prevalence of submicroscopic malaria in pregnant women arriving in non- endemic countries is non negligible ; more data are needed to assess the consequences of submicroscopic malaria on maternal and birth outcomes.

**Running title**

Submicroscopic malaria in pregnancy and migration

### Keywords

### Malaria; submicroscopic; migration; pregnancy

### Title

### Submicroscopic malaria parasitemia and migration: the hidden weight carried by pregnant women.

### Acknowledgments

### This work was funded by the ”Association d’Aide à la Recherche Médicale André Vésale”. The funder provided support in the form of a grant which was used to finance laboratory costs. The funder did not have any additional role in the study design, data collection and analysis, or preparation of the manuscript.

### Biographical Sketch

Dr L. Docquier is a clinical postgraduate trainee in Internal Medecine at the ULB (Université Libre de Bruxelles). She wishes to specialize in tropical diseases research and control, particularly malaria.

Dr C. Taelman is a clinical postgraduate trainee in Gynecology at the ULB (Université Libre de Bruxelles). Her main research area is on malaria.

### Address for Correspondence

### Léa Docquier

### Mailing address : 9 rue Souveraine, 1050 Brussels, Belgium.
